# Assessing the potential for prevention or earlier detection of on-site monitoring findings from randomised controlled trials: further analyses of findings from the prospective TEMPER triggered monitoring study

**DOI:** 10.1101/2020.04.01.20033647

**Authors:** William J Cragg, Caroline Hurley, Victoria Yorke-Edwards, Sally P Stenning

**Author notes:** **Correspondence:** William Cragg, Clinical Trials Research Unit, Leeds Institute of Clinical Trials Research, University of Leeds, Leeds, LS2 9JT, United Kingdom. **Research support:** Medical Research Council (MC_UU_12023/24) The TEMPER study was funded by a grant from Cancer Research UK (C1495/A13305).

## Abstract

**Background/Aims:** Clinical trials should be designed and managed to minimise important errors with potential to compromise patient safety or data integrity, employ monitoring practices that detect and correct important errors quickly, and take robust action to prevent repetition. Regulators highlight the use of risk-based monitoring, making greater use of centralised monitoring and reducing reliance on centre visits. The TEMPER study was a prospective evaluation of triggered monitoring (a risk-based monitoring method), whereby centres are prioritised for visits based on central monitoring results. Conducted in three UK-based randomised cancer treatment trials of investigational medicine products with time-to-event outcomes, it found high levels of serious findings at triggered centre visits but also at visits to matched control centres that, based on central monitoring, were not of concern. Here, we report a detailed review of the serious findings from TEMPER centre visits. We sought to identify feasible, centralised processes which might detect or prevent these findings without a centre visit.

**Methods:** We considered a representative example of each finding type through a three-staged consensus exercise. 1: Three trialists independently reviewed the issues and graded their potential for detection by central monitoring or prevention by some described process, each on a five-point scale. 2. Results of round 1 were shared anonymously and each trialist re-reviewed the list choosing whether or not to change their grading. 3. The three trialists discussed and agreed grades for any issues without consensus, and proposed a feasibility score for each proposed process. In a consultation exercise, the proposed processes were reviewed and rated for feasibility by an invited external trialist group. The primary outcome was the proportion of all major and critical findings theoretically detectable or preventable through a feasible, centralised process.

**Results:** In TEMPER, 312 major or critical findings were identified at 94 visits. These findings comprised 120 distinct issues, for which we proposed 56 different centralised processes. Following independent review of the feasibility of the proposed processes by 87 consultation respondents across different stakeholder groups, we conclude that 306/312 (98%) findings could theoretically be prevented or identified centrally.

**Conclusions:** This work presents a best-case scenario, where a large majority of monitoring findings were deemed theoretically preventable or detectable by central processes. Caveats include the cost of applying all necessary methods, and the resource implications of enhanced central monitoring for both centre and trials unit staff. Of processes not currently deemed feasible, use of electronic health records has the largest potential benefit.

Our results will inform future monitoring plans and emphasise the importance of continued critical review of monitoring processes and outcomes to ensure they remain appropriate.

## Introduction

A well-run clinical trial is designed and managed to minimise damaging errors in conduct.^1^ Monitoring is done to detect important errors in a reasonable timescale and to enable action to prevent repetition. Cumulatively, this helps ensure the safety of trial participants and the integrity of trial results. **Figure 1** shows a suggested relationship between risk, prevention and monitoring.

**Figure 1:**
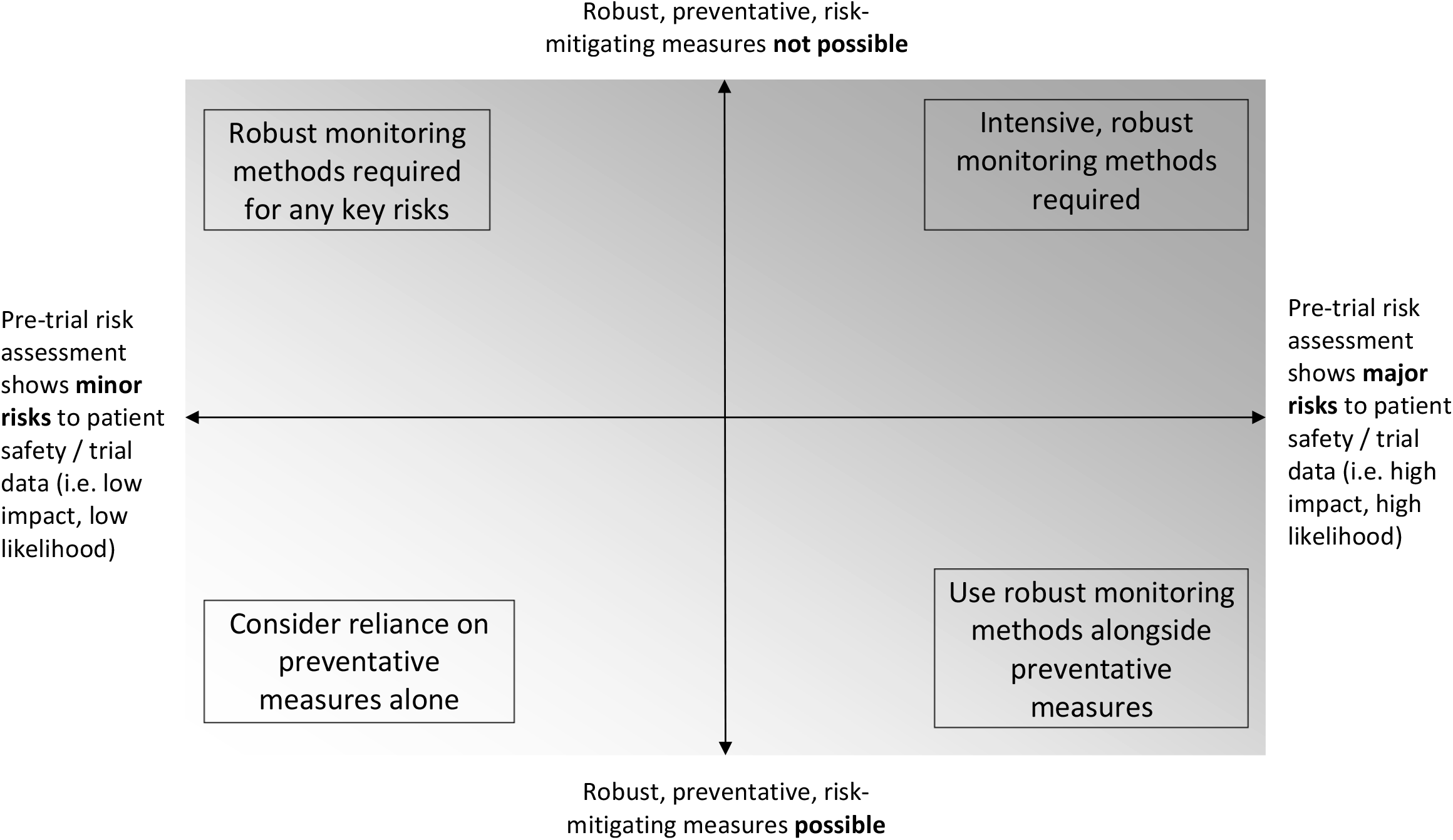
a suggested relationship between preventative measures, monitoring and risk.

Historically, and partly in response to regulatory guidance,^2^ trial monitoring has relied on frequent sponsor visits to trial centres.^3^ These visits have clear benefits in terms of building rapport between sponsor representatives and trial centres, delivering training and achieving trial promotion, as well as the opportunity for in-person review of facilities and source documents.^4^ However, on-site monitoring has some significant limitations. The cost of travel and staff time required for regular centre visits is considerable,^5–7^ and may not be justified given the acknowledged limited benefit of source data verification, a common driver of intensive centre visit strategies.^8–10^ Depending on how frequent visits are, on-site monitoring may detect issues less quickly than central monitoring, i.e. monitoring conducted without centre visits, using data collected from trial centres. Finally, direct access to individual participant source data, while a strength of on-site monitoring, is less useful than central monitoring when looking for trial-wide issues in multicentre trials.

Acknowledging this, current regulatory guidance now encourages risk-based monitoring^11–13^ with greater emphasis on, and even suggested methods for, central monitoring.^14^ There remains, however, a lack of evidence to support different monitoring practices.^3,15,16^ The TEMPER study (TargetEd Monitoring: Prospective Evaluation and Refinement)^17^ assessed whether centrally monitored threshold-based rules – “triggers” – could be used as a means to distinguish clinical trial centres with high or low rates of concerning on-site monitoring findings. In a prospective matched-pair design, centres participating in one of three phase III randomised cancer treatment trials of investigational medicinal products (IMPs) with time-to-event outcomes were prioritised for visits based on the trigger findings. Each triggered centre was matched with one from the same trial that was not triggered, i.e. not of current concern, and both centres were visited. Centre visits were conducted according to each trial’s monitoring plan, but generally included similar activities across all the trials: review of some or all informed consent forms, source data verification and medical notes review for a sample of participants, facility review (including pharmacy), and review of quality and completeness of essential documents. Findings were categorised in terms of seriousness, according to a standardised system, as Critical, Major or Other, using similar definitions to those of UK regulators.^18^ We considered the collection of Critical and Major findings to be those of interest, i.e. ‘errors that matter’.^1^

The full methods and results are reported elsewhere.^17^ TEMPER found that the majority of centres in both triggered visits and matched visits (those without concern) had at least one Major or Critical finding, questioning the efficacy of triggered monitoring as employed in these trials. Here, we report an exploratory review of all the Major and Critical findings reported in TEMPER. We sought to propose centralised monitoring processes or trial process changes which might detect or prevent these findings prior to centre visit. Through this, we aimed to inform and improve future trial conduct by developing an evidence-based (or at least experience-based) central monitoring and quality assurance plan.

## Methods

### Source data: Monitoring Findings from TEMPER study

We used findings from all 94 on-site monitoring visits conducted for TEMPER. There were 312 individual Major or Critical findings (298 Major, 14 Critical); these are summarised in **Table 1**. Some findings had been detected several times within and across centres. In total there were 120 distinct issues; a representative example of each was reviewed as described below. **Figure 2** summarises all the stages of the current study.

**Table 1:** summary of Major and Critical findings at TEMPER monitoring visits.

| Type of finding by Monitoring Report section | Number of findings at all visits (n=94) |  |  | Individual issues <sup>b</sup> |
| --- | --- | --- | --- | --- |
|  | All findings | Major findings<sup>a</sup> | Critical findings<sup>a</sup> |  |
| **Investigator Site File – All** | **6** | **6** | **0** | **5** |
| **Informed consent – All** | **222** | **219** | **3** | **33** |
| *Re-consent (eg failure to obtain re-consent in a timely manner)* | *162* | *162* | *0* | *2* |
| *Original consent (eg missing signatures, missing or in compatible signature dates, incorrect versions used)* | *60* | *57* | *3* | *31* |
| **Pharmacy - All** | **8** | **6** | **2** | **8** |
| **CRF/SDV - All** | **76** | **67** | **9** | **74** |
| *Unreported SAE/notable event* | *25* | *25* | *0* | *25* |
| *Unreported endpoint* | *12* | *12* | *0* | *10* |
| *Source data discrepancy (priority data)* | *19* | *19* | *0* | *19* |
| *Other* | *20* | *11* | *9* | *20* |
| **Total Major and Critical findings** | **312** | **298** | **14** | **120** |

<sup>a</sup> The findings reported from the TEMPER study also included some 'upgrades' to Major and Critical, i.e. groups of findings that collectively indicated a more serious finding. These were not considered relevant outside of the context of the TEMPER study design, and were therefore excluded from this exercise.
<sup>b</sup> Where a finding had occurred multiple times within or across centres, we reviewed only one of each of these duplications as representative of others; these are listed here as 'individual issues' (n=120).

**Figure 2:**
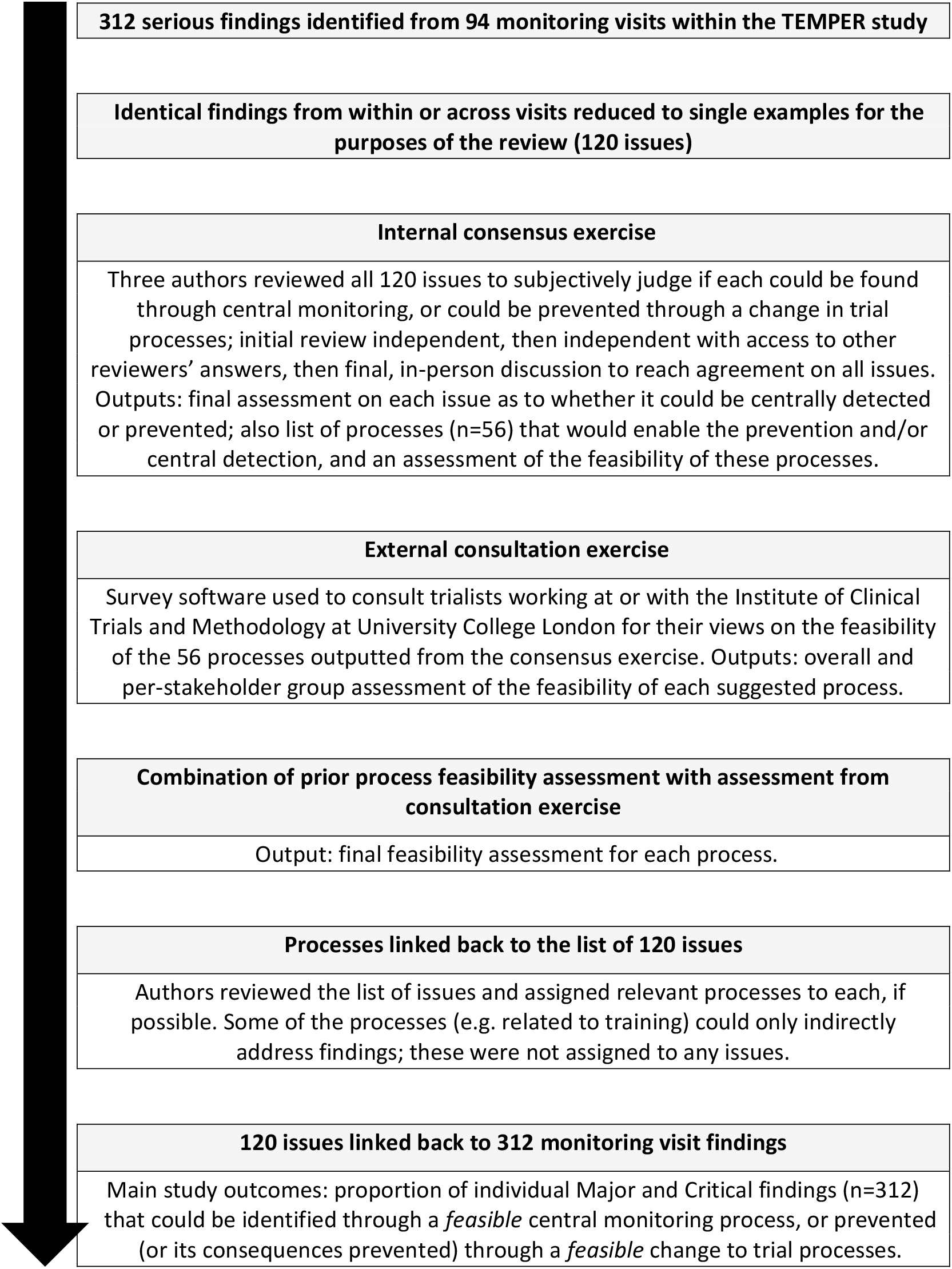
flow diagram describing the current study.

### Initial review and development of suggested processes (consensus exercise)

**Step 1:** independently of each other, three authors (WC, CH, SS) reviewed all 120 issues to consider whether, hypothetically: (1) each issue could be identified through central monitoring; (2) the issue could be prevented, or the consequences prevented, through some change in trial processes; and (3) if the answer to either (1) or (2) was yes, what the central monitoring method or trial process change would be. We considered “prevention” to include both complete prevention of the issue (e.g. process to prevent patient being approached for trial entry if ineligible) and prevention of its consequences (e.g. process to identify erroneously randomised ineligible patient at the time of randomisation and therefore prevent them starting study intervention if not appropriate or safe to do so). Both prevention and central detectability were rated against a five-point scale: 5 = definitely, 4 = possibly, 3 = not sure, 2 = probably not, 1 = definitely not.

**Step 2:** the independent review results were combined into one file. Each reviewer saw their initial responses alongside those of the other, anonymised reviewers and either confirmed or updated their response for each issue.

**Step 3:** reviewers met in-person to agree final results for each issue, using majority votes in cases of any disagreement. The reviewers discussed the potential prevention and central monitoring processes, brainstormed further ideas for these, finalised the process list and agreed a consensus feasibility score for each, using the same five-point scale mentioned above.

### Stakeholder consultation to assess feasibility of suggested processes

We sought independent views on the feasibility of our list of proposed central monitoring and prevention processes through a consultation exercise involving staff at the University College London Institute of Clinical Trials and Methodology, and collaborators working with the Institute’s trials (mainly those working on the three trials involved in TEMPER). We aimed to capture the views of experienced stakeholders both from trial centres (clinicians, research nurses, pharmacists, radiologists) and clinical trials units (trial managers, data managers, trialists and statisticians).

We used Opinio survey software^19^ to develop the consultation exercise. Invitees were not sent any reminders after the initial invite. We asked respondents to review each of our central monitoring and prevention processes (without disclosing our prior feasibility score) and provide a feasibility score from options “Feasible and easy to achieve in current practice” (score 5), “Feasible but expensive or challenging” (4), “Not sure” (3), “Possible but cost or practical issues make it unworkable” (2), “Not possible at present” (1). We also asked respondents whether any of a pre-defined list of challenges might apply to implementing a given process: cost, time for trials unit staff, time for centre staff, logistical issues, legal or regulatory issues, other (specify). There was an opportunity to add free text comments after each of these questions, for each process; these informed our interpretation of process-level and individual results, but are not summarised for this report. We also collected information on respondents’ professional role and their experience with clinical trials and with trial monitoring.

We asked each stakeholder group only about processes we felt were relevant to that group (e.g. we asked pharmacists only about issues of drug management). In cases where many processes (e.g. >25) applied to a single group, the list was split into two and a separate survey link was sent to each half of the whole distribution list.

We carried out data cleaning prior to analysis. 16 respondents had provided free-text comments about the feasibility of particular processes, but no feasibility score; we discussed these and, where we felt the comment was clear, imputed a feasibility score for that respondent. ‘Not sure’ answers were in one of two groups: either respondents understood the proposed process but were not sure of its feasibility, or had said ‘not sure’ to express uncertainty about what the proposed process would entail. Where comments were agreed to unambiguously suggest uncertainty about the nature of the process, we removed the ‘not sure’ response from the final dataset.

No ethical approval was required for the consultation exercise (confirmed using the Health Research Authority’s decision tool^20^).

### Outcomes and analysis

For each process we calculated the median feasibility score from the consultation per stakeholder group, and across all respondents. For comparison with our prior feasibility grade, we grouped these into broader categories: broadly feasible (median score ≥4), broadly not feasible (median score ≤2) and not sure (median score 3). Where consensus and consultation agreed (including across individual stakeholder groups), the broad category was taken as the final answer on the process’ feasibility. Where there was any disagreement in overall or inter-stakeholder medians, we reviewed the consultation data in detail and made a final decision about what broad feasibility category to ascribe. Non-integer, borderline medians, for example a score of 3.5 for one stakeholder group among others ≥4, were taken as disagreement worth further, detailed review. Our general approach was that we would defer to the views of the stakeholders unless we felt they had not understood our description of the proposed process. When respondents suggested a process was feasible only in certain conditions, we categorised as ‘feasible with caveats’.

Returning to the original list of 120 distinct issues, we applied the final feasibility scores to any processes that might address each one. From this, we were able to determine our main overall outcomes, namely the proportion of individual Major and Critical findings (n=312) that could be identified through a feasible central monitoring process, or prevented (or their consequences prevented) through a feasible change to trial processes.

Aside from the survey software, all data collection and analysis took place in Microsoft Excel, and WC managed the data. Independent checks were carried out by SS on a 10% sample of findings and proposed processes respectively, to check feasibility scores pre- and post-consultation.

## Results

In our consensus exercise, we agreed that 114 (95%) of the 120 distinct issues were potentially detectable through central monitoring. A different, mostly overlapping list of 114 (95%) were theoretically preventable through simple changes in trial processes. We proposed 56 processes (or process changes), 43 of which could directly address on-site findings from TEMPER and 13 of which we thought could potentially have an impact, but less directly or without being completely fool-proof, e.g. additional training for trial centre staff.

All 56 processes were included in the consultation exercise, which was run between 20 December 2017 and 26 January 2018. **Table 2** summarises the stakeholder recipients and responders to each process. 87 people completed the exercise, an overall response rate of 19% of those to whom we sent the survey (n=450). We excluded 10 additional responses that provided monitoring experience information only, without answers to the remaining questions. Mean number of years’ experience in clinical trials was 11 (range 2-31). Most respondents had not personally conducted on-site monitoring (54%) or central monitoring (59%), and had worked at a centre that had undergone central or on-site monitoring (55%).

**Table 2:**
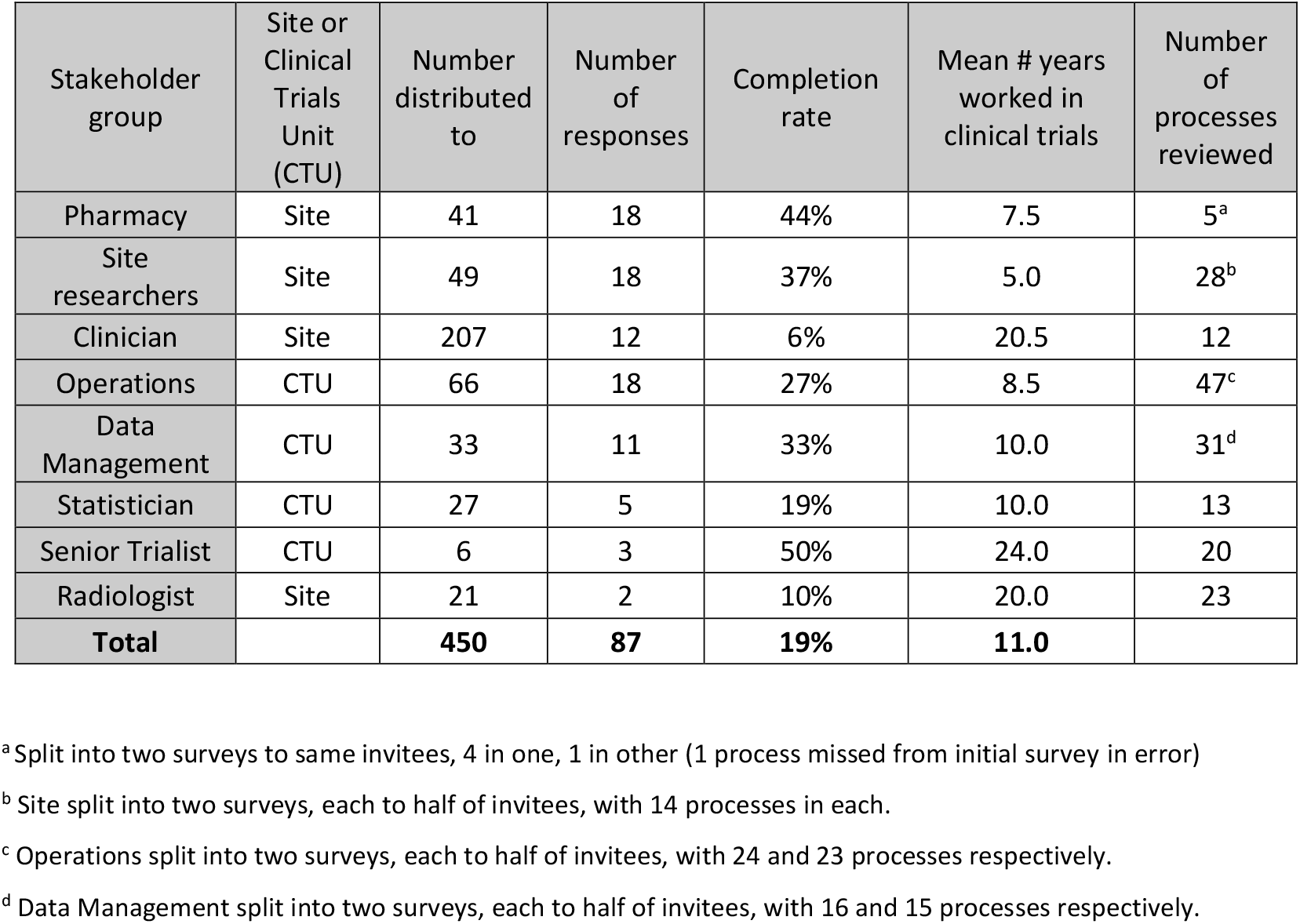
summary of stakeholder groups invited to participate in the consultation exercise, completion rates, clinical trials experience and number of processes reviewed.

For 26/56 of our proposed processes (46%), the median consultation feasibility was in the same broad category (i.e. broadly feasible, broadly not feasible, not sure), across all stakeholder groups, as our prior consensus score. Of the remaining 30/56 processes, 14 were in the same broad category overall, but with differences in some stakeholder groups. After discussing and agreeing final feasibility assessments for these remaining 30, the total 56 were divided into: 43 processes feasible or feasible with a caveat (77%), 5 not feasible (9%) and 8 uncertain (14%).

**Online Table S1a** lists the processes deemed feasible, and the number and proportion of on-site monitoring findings each addresses; **Online Table S1b** lists the processes deemed not currently feasible or of uncertain feasibility, with summarised reasoning for each judgement. **Online Table S1c** summarises the feasibility of the indirect or not fool-proof processes.

Based on the consultation exercise, 304/312 (97%) TEMPER visit findings could be detected through feasible central monitoring methods, and 260/312 (83%) were theoretically preventable, or their consequences preventable, through feasible changes to trial processes. 306/312 (98%) were either centrally detectable, preventable, or both. 256/312 (82%) findings were addressed by more than one suggested process or process change, although this varied across the types of finding (informed consent findings: 222/222, 100%; source data review findings: 21/71, 30%).

**Table 3** lists abridged summaries of the six remaining findings (three each from triggered visits and non-triggered, matched control centre visits) that could not be centrally detected, or prevented, by a feasible process. All were classified Major; no Critical findings remained. All were from Case Report Form checks and source data verification, with no findings remaining from pharmacy, essential document or informed consent form checks. As with the TEMPER study findings as a whole,^17^ there is no indication that these findings would have had any significant impact on the results or interpretation of the trials involved.

**Table 3:** on-site monitoring findings judged not centrally detectable or preventable, following consensus exercise on feasibility of suggested processes.

| Abridged summary of finding | Number of instances | Grading | Reasons why not centrally detectable or preventable |
| --- | --- | --- | --- |
| Incorrectly graded adverse event – reported as CTCAE grade 2 instead of 4 (life-threatening); no serious adverse event reported | 1 | Major | - No simple way to detect this issue without review of source data; if serious adverse event had been submitted, could have cross-checked with those data.<br>- Currently not possible to access and use hospital episode statistics or other electronic health records to detect this.<br>- No obvious fool-proof way to prevent. |
| Unreported serious adverse event; considered 'notable event' in this trial and Case Report Form contained specific question about whether this type of adverse event had occurred, to which answer given had been 'No' | 2 | Major | - Method to centrally identify cases did not work because centre misreported; therefore only way to detect is through review of source data.<br>- Currently not possible to access and use hospital episode statistics or other electronic health records to detect this.<br>- No obvious fool-proof way to prevent. |
| Unreported serious adverse event due to prolongation of hospital stay | 1 | Major | - Prolongation of hospital stay may only be clear from detailed review of medical notes.<br>- Currently not possible to access and use hospital episode statistics or other electronic health records to detect this.<br>- No obvious fool-proof way to prevent. |
| Misreporting of concomitant medication use at randomisation (key baseline data in this trial) | 2 | Major | - Data misreported; not feasible to request source data about concomitant medication to be sent to trials unit for verification, therefore only way to detect is through on-site monitoring.<br>- No obvious fool-proof way to prevent. |

## Discussion

We found that a large majority of important on-site monitoring findings – including all categorised as “Critical” – could theoretically be detected through feasible central monitoring processes or prevented altogether through feasible changes to trial processes. These results corroborate those of the previous, similar work in a different setting.^21^ A large number of findings, especially in informed consent monitoring, could be addressed by more than one process. Exclusion of the 306 “preventable” or “centrally detectable” findings reduces the number of TEMPER monitoring visits with ≥1 Major or Critical finding from 81/94, as in the primary TEMPER study results, to just six (three at triggered visits, three at untriggered).

It is important to note that this represents a best case scenario. Some of the processes will already be in place for some RCTs, including to some degree in the TEMPER trials. We cannot, therefore, easily say how many potential on-site monitoring findings were already successfully avoided through use of these processes in these trials. We did not aim to prove that the TEMPER on-site monitoring findings would definitely have been found earlier through our suggested processes, only to use the findings to develop a more comprehensive plan for future trials. Nonetheless, this may imply that, as currently used, some processes may be insufficient to prevent or centrally identify all issues, or that they cannot be – or have not so far been - implemented consistently across centres and time. Furthermore, an unavoidable limitation of many central monitoring processes is their reliance on good quality and timely reporting from centres.

An increased focus on central rather than on-site monitoring may have implications for resourcing. From free-text responses to our consultation exercise (data not shown), it is clear some centre research staff already feel that some central monitoring processes make large demands of their time, without adequate resourcing or recognition (as noted by others^22^). Central monitoring also displaces resource within the trials unit from dedicated on-site monitors towards the database programmers and statisticians responsible for developing reports and reviewing data, and trial management staff responsible for following up highlighted issues.^23^ Resolving these resourcing questions, perhaps partly through different financial arrangements with centres in trials that rely more on central monitoring, could be a necessary precursor to a more widespread adoption of central monitoring methods.

Our Major and Critical findings were dominated by errors relating to informed consent, because of (1) the importance of consent forms in clinical trial legislation, and therefore also in our monitoring plans and findings categorisation scheme, (2) the relative frequency of errors (not apparently atypical^15,24,25^), (3) the monitoring approach: only one of the three TEMPER trials centrally monitored consent forms, and then not as a pre-randomisation check, and (4) the relatively frequent re-consent requests in these trials, as reported in the main TEMPER report.^17^ We believe (with the support of our consultation) all our findings relating to initial informed consent forms are detectable centrally, at the point of randomisation, thereby preventing patients starting treatment if there are any issues. By the time of TEMPER’s main publication in 2018, a number of RCTs at the University College London Institute of Clinical Trials and Methodology had started to carry out routine, pre-randomisation central consent form monitoring, whereby a copy of the completed form is checked at the trials unit before being destroyed securely. This may be less feasible in trials with short lead-up times, such as some trials in emergency medicine. Future developments in electronically documenting consent^26,27^ may be useful in resolving many issues we currently face with paper-based forms.

Of the remaining findings, a large proportion related to patient eligibility (recognised as a problem in other trials^28^), unreported Serious Adverse Events and unreported time-to-event endpoints (in these RCTs, death or cancer progression). Processes to check eligibility prior to randomisation include collection of more detailed data on trial forms to verify eligibility (e.g. blood results) rather than just tick-box confirmation that a patient meets each eligibility criterion, and collecting pseudonymised copies of source data to verify key aspects of eligibility. Our consultation confirmed that both of these are possible, although perhaps not practical in all trials. Some consultation respondents also voiced concern about the availability of clinical expertise required to review medical data before randomisation; it may therefore be best to limit this to objective assessments (e.g. checking blood results are within range) rather than more complex assessments of, for example, scan reports.

Timely reporting of Serious Adverse Events is fundamental to adhering to regulatory requirements for RCTs, and sponsors must have processes in place to ensure all events are received from trial centres. Our consultation exercise reported uncertainty about the current possibility and timeliness of using routinely-collected health data in the UK (e.g. Health Episode Statistics data^29^) to identify unreported events. For now, our best suggestion is therefore to collect regular data to help ascertain if any Serious Adverse Events may have taken place, such as whether there have been any in-patient hospitalisations or additional treatments since the last follow-up visit. We also suggest that better informing trial participants about safety reporting requirements, particularly where emergency admissions may take place at a non-trial hospital, may improve reporting rates.

Unreported deaths in trials with survival-based outcome measures can be relatively simple to detect centrally by, for example, closely following-up cases where there is no data at the trials unit about a given patient for a long time. It is also feasible to obtain data on patient deaths through national registries, though challenges may remain regarding associated costs and the timeliness of available data. Unreported disease progression can be more difficult to centrally detect, depending on how much additional indicative data is collected. There may be scope for using routinely-collected health data to look for changes in patient treatment that may indicate disease progression, however our consultation respondents felt we could not say this was feasible in the UK at present.

In the list of processes considered not feasible in our consultation, processes involving use of routinely-collected health data were among those with the largest *potential* impact on monitoring. Although there are some reported examples of remote access to individual patient records for source data verification,^30–32^ there are currently substantial legal and information governance barriers to this becoming routine, certainly in the UK. The feasibility of regular, timely access to linked electronic health record data for research purposes is easier to envisage, and could facilitate identification of unreported (serious) adverse events, trial outcome data and verification of, for example, health economics data. The suitability and availability of these data for complex, time-sensitive purposes is yet to be proven, however.^33,34^

We acknowledge several limitations not already mentioned. The central monitoring processes we propose may be most suitable for trials like those included in TEMPER, which were late-phase trials with already-licensed IMPs posing moderate risk to trial participants. A higher degree of reliance on central monitoring may be less appropriate for higher-risk trials. The feasibility of our monitoring and prevention processes was confirmed not through a broad survey of trialists, but through a convenience sample of staff and collaborators, mainly working on trials of cancer treatments, at the University College London Institute of Clinical Trials and Methodology in the UK. The consultation exercise had a relatively low response rate. Our results may therefore not be generalizable to different settings.

We asked our consultation respondents to comment on each suggested process in isolation, and to implement all processes in combination would be a significant undertaking. This again highlights the importance of properly resourcing all central monitoring activity. Finally, we should acknowledge that we could implement all these suggested processes in a subsequent trial in a different setting and find different, important findings at trial centres. Comparison with our earlier work^21^ supports this to some extent.

We recommend that trialists review the most common errors in their trials and give due consideration to implementing the central processes – including, but not limited to, those reported in this work – that could detect or prevent the majority of serious issues, alongside processes to address additional, trial-specific risks. There is scope for more formal evaluation of our resulting monitoring plan, and generation of additional data on efficacy and costs, both direct and indirect.

In conclusion, we recommend the process we have explored here of systematically using the results of monitoring to induce continual improvement of trial processes should be a routine, ongoing exercise, as it will ultimately lead to more robust data, safer participants and better trials. Standardised, systematic recording of data about clinical trial monitoring may be a necessary precursor to this, to facilitate review and assessment of trends within and across trials.

## Data Availability

The data supporting this work are available on reasonable request. Please contact the corresponding author in the first instance.
The underlying data were collected as part of the TEMPER study. Please refer to the main study publication for details of how to access these data (Stenning et al, Clinical Trials 15:6(600-609), doi: 10.1177/1740774518793379)

## Acknowledgements

We would like to acknowledge the support of those who helped this work by participating in the consultation exercise, and of Matthew R Sydes, Sharon B Love and Mahesh KB Parmar at Medical Research Council Clinical Trials Unit at University College London for their reviews of draft manuscripts. We also thank the wider TEMPER study team for their support, the trial teams whose trials took part in TEMPER and the site staff whose visits were included in TEMPER.

## Conflicts of interest

The Authors declare that there are no conflicts of interest.

## Funding

The TEMPER study was funded by a grant from Cancer Research UK (grant C1495/A13305) from the Population Research Committee); additional support for the present work was provided by the Medical Research Council London Hub for Trial Methodology Research (MC_UU_12023/24).

## Supplementary information

- Supplementary tables S1a-c
- Consultation exercise text

## Summary of supplementary information

1. **Online Tables S1**
  - **Online Table S1a**: all processes agreed after the consensus exercise to be feasible or feasible if adjusted in specific ways, and the number and proportion of findings each addresses, overall and by subtype.
  - **Online Table S1b:** all processes agreed after the consensus exercise to be not feasible or of uncertain feasibility, and the number and proportion of findings each addresses, overall and by subtype.
  - **Online Table S1c:** processes that could address on-site monitoring findings, but indirectly or without being completely fool-proof, with feasibility rating and, where applicable, suggested adjustments to improve feasibility or reasons why infeasible.
2. **Consultation exercise text**

## Notes

### Competing Interest Statement

The authors have declared no competing interest.

